# Intersectional Socioeconomic Inequalities in Maternal and Reproductive Health Outcomes Among Cambodian Women: A Secondary Analysis of the 2021–22 Demographic and Health Survey

**DOI:** 10.64898/2025.12.20.25342731

**Authors:** Yem Sokha, Kem Sokunthea, Nov Sreyroth, Meas Moniroth, Tun Sreypeov

## Abstract

**Background:** Health inequalities in low- and middle-income countries (LMICs) often reflect compounding social disadvantages. Intersectionality theory emphasizes that multiple axes of disadvantage can interact to shape health experiences in ways not captured by single-factor analyses (Crenshaw, 1989, 1991). Understanding how poverty and low education combine to affect maternal and reproductive health is essential for advancing universal health coverage and “leaving no one behind” under the Sustainable Development Goals (United Nations, 2015; World Health Organization, 2023). This study examined intersectional socioeconomic inequalities in maternal and reproductive health outcomes among Cambodian women.

**Methods:** We conducted a secondary analysis of women aged 15–49 years interviewed in the Cambodia Demographic and Health Survey (CDHS) 2021–22, a nationally representative household survey implemented by the National Institute of Statistics (NIS) in collaboration with the Ministry of Health, with technical assistance from ICF (National Institute of Statistics et al., 2022). Women were classified into six intersectional socioeconomic status (SES) groups by cross-classifying household wealth (poor/middle/rich) and education (no/primary vs. secondary+). Outcomes included antenatal care (ANC) ≥4 visits, facility delivery, skilled birth attendance, modern contraception use, unmet need for family planning, and teenage childbearing. Multivariable logistic regression estimated adjusted odds ratios (aORs) with 95% confidence intervals (CIs), accounting for the complex survey design (National Institute of Statistics et al., 2022).

**Results:** Among 6,968 women with recent births, 82% attended ≥4 ANC visits and 96% delivered in facilities, but only 20% had skilled birth attendance. Compared with women with both high wealth and secondary+ education (reference), women with poor wealth and no/primary education had substantially lower odds of ≥4 ANC visits (aOR = 0.33, 95% CI [0.23, 0.48]), facility delivery (aOR = 0.29, 95% CI [0.13, 0.63]), and skilled birth attendance (aOR = 0.20, 95% CI [0.14, 0.27]). For reproductive health, poor/low-education women had higher odds of modern contraception use (aOR = 1.24, 95% CI [1.06, 1.43]) and no difference in unmet need (aOR = 0.99, 95% CI [0.85, 1.16]). Teenage childbearing showed strong educational gradients (35% in poor/low-education vs. 16% in rich/high-education).

**Conclusions:** Intersectional socioeconomic disadvantage creates compounding barriers to quality maternal healthcare in Cambodia. Despite near-universal facility delivery, skilled attendance—defined in global monitoring as births attended by trained health personnel such as doctors, nurses, or midwives—remains highly unequal and concentrated among advantaged groups (United Nations Statistics Division, 2023). Policies should prioritize equitable quality of delivery care, target multiply disadvantaged women, strengthen rural health workforce distribution, and invest in girls’ secondary education as a long-term maternal health strategy (National Institute of Statistics et al., 2015; World Health Organization, 2023).

## Introduction

Health inequalities in LMICs reflect overlapping social and structural determinants, including poverty, education, gender, and geography, that shape access to and quality of care. While national indicators may improve, within-country disparities can persist—challenging progress toward universal health coverage and SDG 3 (United Nations, 2015; World Health Organization, 2023). Cambodia has experienced major improvements in maternal health service use over time, including increases in skilled delivery assistance: preliminary DHS reporting noted that births delivered by a health professional rose from **44% (CDHS 2005)** to **71% (CDHS 2010)** (National Institute of Statistics et al., 2010), and the CDHS 2014 reported that **89% of births** in the five years preceding the survey were assisted by a trained health professional (doctor, nurse, or midwife) (National Institute of Statistics et al., 2015). However, progress in coverage can mask inequities in who receives *high-quality* care and whether essential components of safe childbirth are equitably available. Traditional inequality analyses often examine wealth or education separately, but intersectionality theory argues that social disadvantages interact, producing experiences not reducible to single dimensions (Crenshaw, 1989, 1991). In Southeast Asia, few studies have explicitly evaluated how combined socioeconomic disadvantages shape maternal and reproductive outcomes. This study addresses this gap by applying an intersectional wealth × education framework to CDHS 2021–22 data to examine inequalities across antenatal care utilization, delivery care, family planning indicators, and teenage childbearing (National Institute of Statistics et al., 2022).

## Methods

We conducted a cross-sectional secondary analysis of the CDHS 2021–22, a nationally representative household survey implemented by the National Institute of Statistics in collaboration with the Ministry of Health, with technical assistance from ICF (National Institute of Statistics et al., 2022). The survey used a two-stage stratified design based on the 2019 General Population Census sampling frame: **709 clusters** were selected in the first stage, and **30 households per cluster** were systematically selected in the second stage for a total sample of **21**,**270 households** (National Institute of Statistics et al., 2022). All women aged 15–49 who were usual residents or visitors present the night before the survey were eligible for interview (National Institute of Statistics et al., 2022). We created an intersectional SES exposure by cross-classifying household wealth and education. Household wealth was measured using the DHS wealth index derived from household assets and characteristics using principal components analysis (Rutstein & Johnson, 2004); we collapsed wealth quintiles into poor (Q1–Q2), middle (Q3), and rich (Q4–Q5). Education was dichotomized as no/primary versus secondary or higher. Cross-classification produced six mutually exclusive SES groups, with rich + secondary+ as the reference. Maternal health outcomes among women with births in the past five years (most recent birth) included ANC ≥4 visits, facility delivery, and skilled birth attendance (doctor, nurse, or midwife). Reproductive outcomes among all women included modern contraception use, unmet need for family planning, and teenage childbearing (first birth <20 years among women who ever gave birth). Models adjusted for age group, urban/rural residence, and province; contraception and unmet need models additionally adjusted for marital status. All analyses accounted for survey weights, clustering, and stratification (National Institute of Statistics et al., 2022). We estimated adjusted odds ratios (aORs) and 95% CIs using multivariable logistic regression and used the subpopulation approach for restricted analyses to ensure appropriate variance estimation. Statistical significance was assessed at α =.05 (two-tailed).

## Ethical Considerations

The CDHS protocol received ethical approval, and all participants provided informed consent; this secondary analysis used de-identified publicly available DHS data and did not require additional ethical approval (National Institute of Statistics et al., 2022).

## Results

**Skilled birth attendance** remained low despite near-universal facility delivery. Nationally, only 20.4% of women reported being attended during delivery by a doctor, nurse, or midwife, consistent with the global monitoring concept of “skilled health personnel” (United Nations Statistics Division, 2023). This outcome showed the steepest intersectional gradient: only 12.3% of poor women with no/primary education had skilled attendance compared with 45.3% among rich women with secondary+ education, a 33 percentage-point gap (Table 2). In adjusted analyses (Table 3, Model 3), all SES groups had significantly lower odds of skilled attendance than the most advantaged reference group (rich + secondary+), including poor/no-primary (aOR = 0.20, 95% CI [0.14, 0.27]), poor/secondary+ (aOR = 0.34, 95% CI [0.25, 0.48]), middle/no-primary (aOR = 0.26, 95% CI [0.18, 0.40]), middle/secondary+ (aOR = 0.49, 95% CI [0.35, 0.68]), and rich/no-primary (aOR = 0.46, 95% CI [0.34, 0.61]) (all p <.001). Notably, even relatively advantaged non-reference groups had less than half the odds of skilled attendance, suggesting a strong concentration of skilled delivery care among the most socioeconomically advantaged women.

**Table 1.**
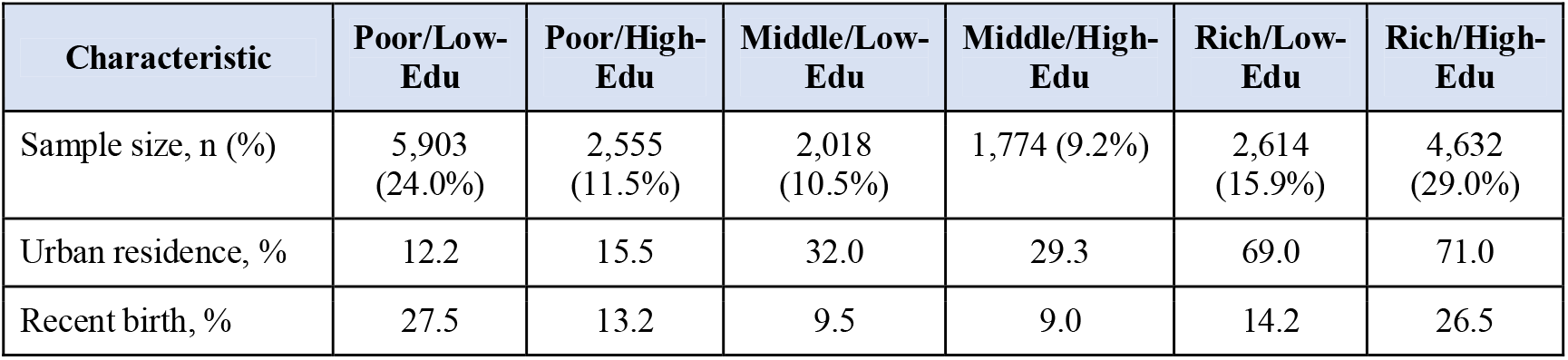
Sample Characteristics by Intersectional SES Category.

**Table 2.**
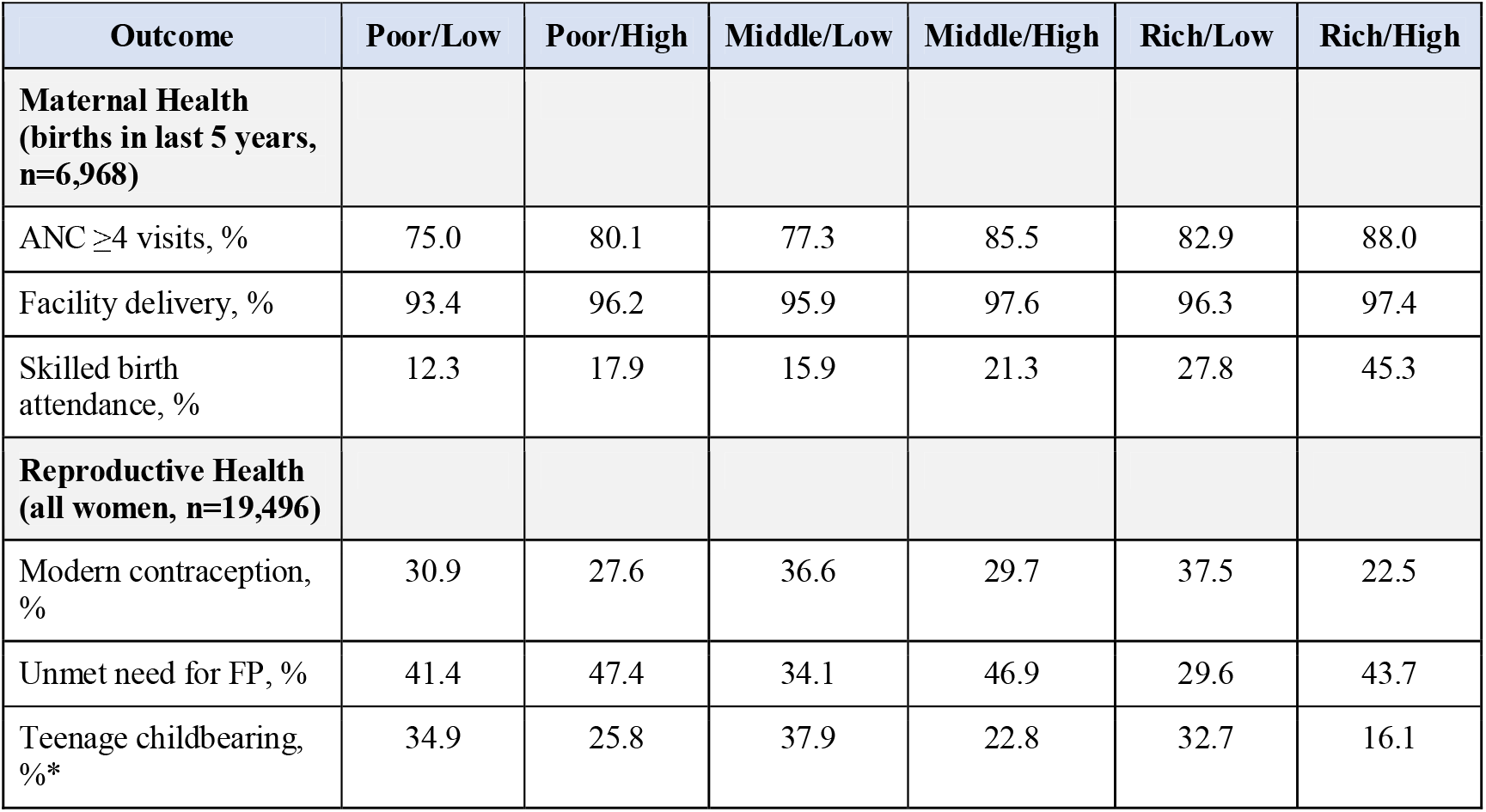
Outcome Prevalence by Intersectional SES Category.

**Table 3.**
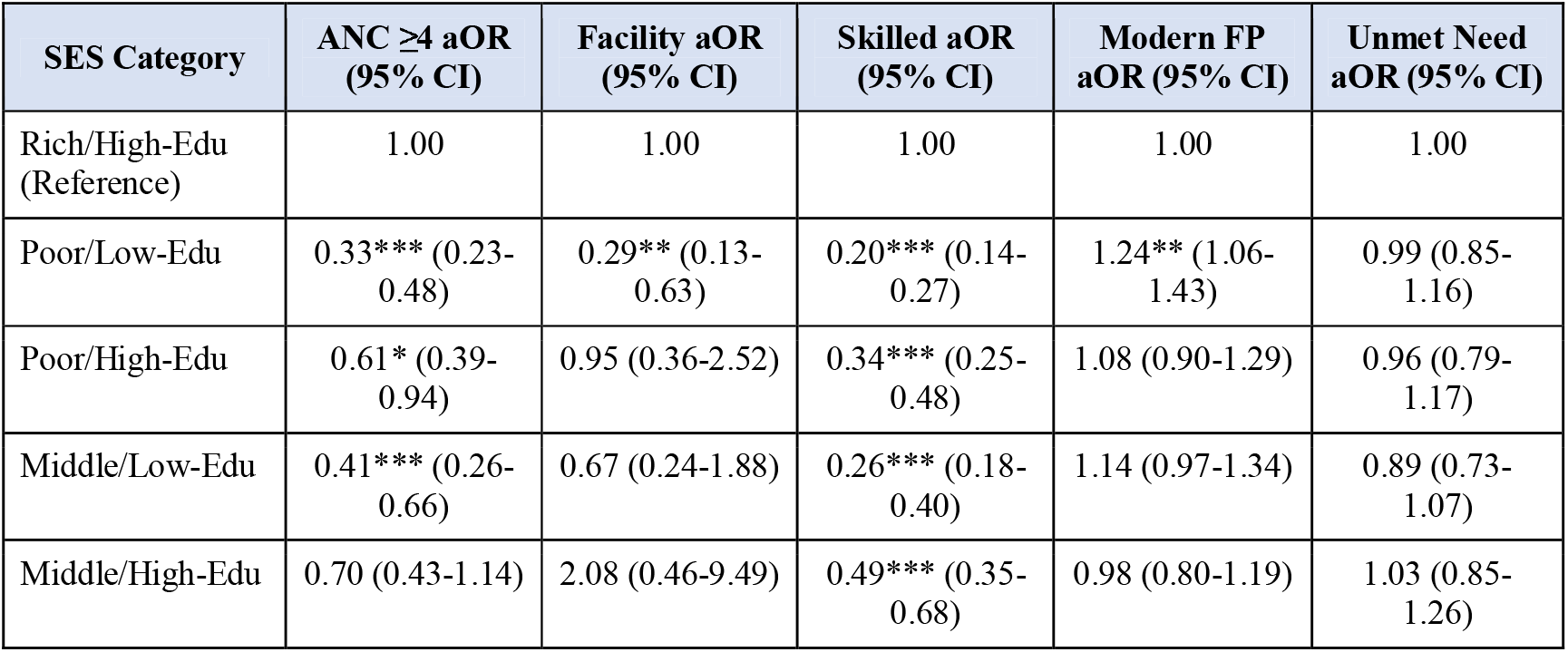
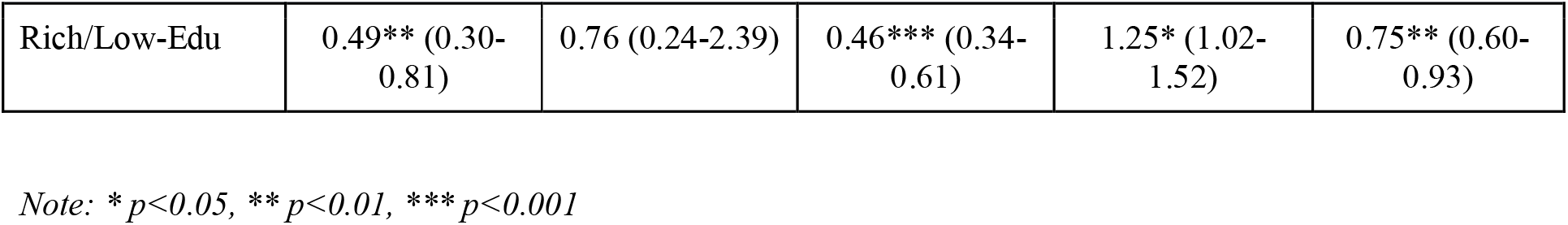
Adjusted Odds Ratios for Maternal and Reproductive Health Outcomes.

## Discussion

This study applied an intersectional framework to examine socioeconomic inequalities in maternal and reproductive health outcomes among Cambodian women, revealing that combined disadvantages of poverty and low education create compounding barriers to high-quality maternal healthcare. Our findings demonstrate that while Cambodia has achieved near-universal facility delivery coverage (96%), substantial inequities persist in the quality dimension of care, particularly skilled birth attendance. Only 20% of women nationally received skilled attendance during delivery, with stark gradients across intersectional SES groups: women experiencing both poverty and low education had 80% lower odds of skilled attendance compared with their wealthy, educated counterparts. This pattern suggests that health system improvements have not equitably reached all population subgroups, and that aggregate national indicators can mask profound within-country disparities (Victora et al., 2003).

The disconnect between high facility delivery rates and low skilled attendance rates warrants particular attention. This finding indicates that many women in Cambodia deliver in health facilities without access to doctors, nurses, or midwives—the cadres recognized internationally as skilled birth attendants (United Nations Statistics Division, 2023). Several explanations merit consideration. First, rural health facilities may lack adequate staffing with qualified personnel, relying instead on auxiliary nurses or other cadres not classified as skilled under global monitoring standards (Ir et al., 2015). Second, geographic maldistribution of the health workforce concentrates skilled providers in urban areas and provincial hospitals, leaving rural commune health centers underserved (Dawson et al., 2016). Third, socioeconomic barriers may prevent disadvantaged women from accessing higher-level facilities where skilled personnel are more reliably present, even when they deliver within the formal health system. This quality-coverage gap has important implications for maternal mortality reduction, as skilled attendance during delivery is essential for timely recognition and management of obstetric complications (World Health Organization, 2023).

The educational gradient observed across multiple outcomes aligns with extensive evidence that women’s education—particularly secondary schooling—is among the most powerful determinants of maternal health behaviors and outcomes (Karlsen et al., 2011). Education may operate through multiple pathways: enhanced health literacy and knowledge of danger signs, greater autonomy in healthcare decision-making, increased household resources and social capital, and improved ability to navigate health systems (Karlsen et al., 2011). Our finding that teenage childbearing was more than twice as prevalent among poor women with low education (35%) compared with wealthy, educated women (16%) underscores the importance of keeping girls in school as a dual investment in educational attainment and reproductive health. Early childbearing is associated with elevated risks of maternal morbidity and mortality, adverse birth outcomes, and intergenerational cycles of poverty and poor health (Ganchimeg et al., 2014).

The intersectional approach employed in this study offers methodological and conceptual advantages over traditional single-axis inequality analyses. By simultaneously considering wealth and education, we identified groups experiencing multiplicative disadvantage that would remain hidden in stratification by either dimension alone (Bauer, 2014). For instance, poor women with secondary education and wealthy women with low education occupy distinct positions in the intersectional matrix and face different configurations of barriers and facilitators. Intersectionality theory, originally developed to theorize the experiences of Black women facing compounded race and gender discrimination (Crenshaw, 1989, 1991), provides a valuable lens for understanding how structural inequalities in LMICs are shaped by overlapping systems of social stratification. Future health equity research should expand intersectional analyses to include additional axes of disadvantage such as ethnicity, rural/urban residence, and subnational region, which are known to interact with SES in shaping health outcomes in Cambodia and other Southeast Asian contexts (Houweling et al., 2019).

The unexpected finding that poor women with low education had higher odds of modern contraception use compared with the reference group requires careful interpretation. This may reflect successful targeting of family planning programs toward disadvantaged populations, Cambodia’s long-standing commitment to expanding contraceptive access through community-based distribution and social marketing (National Institute of Statistics et al., 2015), or differential fertility preferences across SES groups. Alternatively, this pattern could indicate limited access to alternative contraceptive methods or facility-based services among advantaged women who may prefer provider-dependent methods. The absence of significant SES differences in unmet need for family planning suggests relatively equitable access to contraception, which stands in contrast to the steep gradients observed for maternal health services and likely reflects different programmatic histories and delivery strategies.

Our findings align with and extend previous research on maternal health inequalities in Cambodia and comparable LMICs. Earlier analyses of CDHS data documented wealth-based and education-based gradients in facility delivery and skilled attendance (National Institute of Statistics et al., 2015), but the intersectional framework employed here reveals that these gradients are mutually reinforcing rather than independent. Studies from other South and Southeast Asian countries have similarly documented that women facing multiple disadvantages experience the greatest barriers to quality maternal healthcare (Houweling et al., 2019; Sahoo & Madhanraj, 2021). The concentration of skilled delivery care among advantaged groups observed in our data reflects broader patterns of health system inequity in which higher-quality services and more specialized providers are disproportionately accessible to urban, wealthy, and educated populations (Victora et al., 2003).

## Limitations

This study has several limitations that should be considered when interpreting findings. First, the cross-sectional design precludes causal inference; observed associations between intersectional SES and health outcomes may reflect unmeasured confounding, reverse causation, or selection effects. For example, teenage childbearing may lead to truncated education and poverty rather than solely resulting from these conditions. Longitudinal data would be needed to clarify temporal relationships and mechanisms. Second, we relied on self-reported outcomes, which may be subject to recall bias, social desirability bias, or misclassification. The five-year recall period for maternal health outcomes, while standard in DHS surveys, may introduce error, particularly for women with multiple births (National Institute of Statistics et al., 2022). Validation studies comparing self-reported skilled attendance with facility records would strengthen confidence in these estimates.

Third, our intersectional framework included only two dimensions of stratification— household wealth and education—due to sample size considerations and analytical tractability. Other important axes of inequality, including ethnicity, rural/urban residence, subnational region, age, and marital status, were controlled as covariates but not incorporated into the intersectional classification. This simplified approach may miss important subgroup disparities. For instance, ethnic minority women in Cambodia face compounded disadvantages in healthcare access that intersect with poverty and education in complex ways (Ensor et al., 2017), but we were unable to examine ethnic intersectionality due to limited sample sizes in minority subgroups. Future research should explore higher-order intersectional classifications where data permit and consider qualitative methods to understand lived experiences of multiply marginalized women.

Fourth, the DHS wealth index, while widely used and validated for cross-country comparisons, has recognized limitations (Rutstein & Johnson, 2004). It is a relative measure that ranks households within a country but does not capture absolute poverty or income. The index emphasizes asset ownership and housing characteristics that may be less relevant for capturing economic well-being in certain contexts. Additionally, wealth is measured at the household level and may not reflect women’s individual economic resources or intra-household inequalities in resource allocation. Complementary analyses using consumption expenditure data or multidimensional poverty measures could provide additional insights.

Fifth, our measure of skilled birth attendance relied on respondents’ reports of the provider type present at delivery (doctor, nurse, or midwife), which may be subject to misclassification if respondents were uncertain about provider qualifications or if local terminology for health worker cadres is ambiguous. The low prevalence of skilled attendance (20%) relative to facility delivery (96%) suggests either substantial underreporting, actual gaps in skilled provider availability, or both. Objective measures from facility assessments or direct observation would provide more reliable estimates of skilled provider presence and quality of care. Finally, we could not assess quality dimensions beyond provider type, such as adherence to evidence-based practices, respectful care, or availability of essential supplies and equipment—all of which influence maternal health outcomes and may also vary systematically by women’s SES (Kruk et al., 2018).

## Policy Implications

These findings suggest several policy priorities. First, Cambodia’s maternal health strategy should emphasize **quality over coverage** by moving beyond facility delivery as the primary performance metric and ensuring that every delivery has access to skilled attendance—an SDG-tracked indicator of maternal health system functioning (United Nations Statistics Division, 2023). This will require recruiting, deploying, and retaining skilled providers in rural areas through improved incentives, housing, continuing professional development, and clearer career pathways, alongside stronger facility staffing standards that ensure skilled personnel availability (World Health Organization, 2023). Second, programs should **explicitly target multiply disadvantaged women**—particularly those who are poor and have limited education—through strengthened community outreach, maternity waiting homes, transport vouchers, conditional cash transfers, and accompaniment models, with geographic focus on rural districts (Crenshaw, 1989, 1991). Third, investments that keep girls in **secondary education** should be framed as long-term health investments because education is strongly associated with safer maternal health behaviors and service uptake (National Institute of Statistics et al., 2015). Fourth, routine monitoring should **disaggregate outcomes by intersectional SES (wealth × education)**, rather than only single-axis stratification, to reveal hidden inequities and guide resource allocation toward those at highest risk (Crenshaw, 1991; World Health Organization, 2023). Finally, because urban–rural gaps can compound socioeconomic disadvantage, policies should address **geographic inequity** through redistribution mechanisms (e.g., rural service requirements or rural premiums) and targeted strengthening of rural maternity services.

## Data Availability

https://dhsprogram.com/data/

## Author Contributions

YS conceptualized the study, designed the methodology, conducted the formal analysis, and wrote the original draft. KS contributed to methodology development and data curation. NS contributed to literature review and validation. MM assisted with data visualization and writing review. TS provided supervision and critical review of the manuscript. All authors reviewed and approved the final manuscript.

## Funding

This research received no specific grant from any funding agency in the public, commercial, or not-for-profit sectors.

## Conflicts of Interest

The authors declare no conflicts of interest.

## Data Availability Statement

The data used in this study are publicly available from the DHS Program at https://dhsprogram.com. Access requires registration and approval from the DHS Program.

## Notes

### Competing Interest Statement

The authors have declared no competing interest.

### Funding Statement

This study did not receive any funding

## References

Bauer, G. R. (2014). Incorporating intersectionality theory into population health research methodology: Challenges and the potential to advance health equity. Social Science & Medicine, 110, 10–17. 10.1016/j.socscimed.2014.03.022

Crenshaw, K. (1989). Demarginalizing the intersection of race and sex: A Black feminist critique of antidiscrimination doctrine, feminist theory and antiracist politics. University of Chicago Legal Forum, 1989(1), 139–167. https://chicagounbound.uchicago.edu/uclf/vol1989/iss1/8/

Crenshaw, K. (1991). Mapping the margins: Intersectionality, identity politics, and violence against women of color. Stanford Law Review, 43(6), 1241–1299. 10.2307/1229039

Dawson, A. J., Nkowane, A. M., & Whelan, A. (2016). Approaches to improving the contribution of the nursing and midwifery workforce to increasing universal access to primary health care for vulnerable populations: A systematic review. Human Resources for Health, 13(1), Article 97. 10.1186/s12960-015-0096-1

Ensor, T., Chhun, C., Kimsun, T., McPake, B., & Edoka, I. (2017). Impact of health financing policies in Cambodia: A 20 year experience. Social Science & Medicine, 177, 118–126. 10.1016/j.socscimed.2017.01.034

Ganchimeg, T., Ota, E., Morisaki, N., Laopaiboon, M., Lumbiganon, P., Zhang, J., Yamdamsuren, B., Temmerman, M., Say, L., Tunçalp, Ö., Vogel, J. P., Souza, J. P., & Mori, R. (2014). Pregnancy and childbirth outcomes among adolescent mothers: A World Health Organization multicountry study. BJOG: An International Journal of Obstetrics & Gynaecology, 121(Suppl. 1), 40–48. 10.1111/1471-0528.12630

Houweling, T. A. J., Tripathy, P., Nair, N., Rath, S., Rath, S., Gope, R., Sinha, R., Bajpai, A., Prost, A., & Costello, A. (2019). The equity impact of participatory women’s groups to reduce neonatal mortality in India: Secondary analysis of a cluster-randomised trial. International Journal for Equity in Health, 18(1), Article 170. 10.1186/s12939-019-1075-4

Ir, P., Horemans, D., Souk, N., & Van Damme, W. (2015). Using targeted vouchers and health equity funds to improve access to skilled birth attendants for poor women: A case study in three rural health districts in Cambodia. BMC Pregnancy and Childbirth, 10(1), Article 1. 10.1186/1471-2393-10-1

Karlsen, S., Say, L., Souza, J. P., Hogue, C. J., Calles, D. L., Gülmezoglu, A. M., & Raine, R. (2011). The relationship between maternal education and mortality among women giving birth in health care institutions: Analysis of the cross sectional WHO Global Survey on Maternal and Perinatal Health. BMC Public Health, 11(1), Article 606. 10.1186/1471-2458-11-606

Kruk, M. E., Gage, A. D., Arsenault, C., Jordan, K., Leslie, H. H., Roder-DeWan, S., Adeyi, O., Barker, P., Daelmans, B., Doubova, S. V., English, M., Elorrio, E. G., Guanais, F., Gureje, O., Hirschhorn, L. R., Jiang, L., Kelley, E., Lemango, E. T., Liljestrand, J., … Pate, M. (2018). High-quality health systems in the Sustainable Development Goals era: Time for a revolution. The Lancet Global Health, 6(11), e1196–e1252. 10.1016/S2214-109X(18)30386-3

National Institute of Statistics, Directorate General for Health, & ICF Macro. (2010). Cambodia Demographic and Health Survey 2010: Preliminary report. National Institute of Statistics, Directorate General for Health, and ICF Macro. https://dhsprogram.com/pubs/pdf/PR16/PR16.pdf

National Institute of Statistics, Directorate General for Health, & ICF International. (2015). Cambodia Demographic and Health Survey 2014. National Institute of Statistics, Directorate General for Health, and ICF International. https://dhsprogram.com/pubs/pdf/FR312/FR312.pdf

National Institute of Statistics, Ministry of Health, & ICF. (2022). Cambodia Demographic and Health Survey 2021–22: Key indicators report. National Institute of Statistics, Ministry of Health, and ICF. https://dhsprogram.com/pubs/pdf/PR130/PR130.pdf

Rutstein, S. O., & Johnson, K. (2004). The DHS wealth index (DHS Comparative Reports No. 6). ORC Macro. https://dhsprogram.com/pubs/pdf/CR6/CR6.pdf

Sahoo, H., & Madhanraj, K. (2021). Demographic and socio-economic determinants of maternal health care service utilization in India: A comparative analysis across EAG and non-EAG states. Clinical Epidemiology and Global Health, 12, Article 100832. 10.1016/j.cegh.2021.100832

United Nations. (2015). Transforming our world: The 2030 agenda for sustainable development (A/RES/70/1). https://sdgs.un.org/2030agenda

United Nations Statistics Division. (2023). SDG indicator 3.1.2 metadata: Proportion of births attended by skilled health personnel. https://unstats.un.org/sdgs/metadata/files/Metadata-03-01-02.pdf

Victora, C. G., Wagstaff, A., Schellenberg, J. A., Gwatkin, D., Claeson, M., & Habicht, J. P. (2003). Applying an equity lens to child health and mortality: More of the same is not enough. The Lancet, 362(9379), 233–241. 10.1016/S0140-6736(03)13917-7

World Health Organization. (2023). Universal health coverage. https://www.who.int/health-topics/universal-health-coverage

